# Rapid screening for variants of concern in routine SARS-CoV-2 PCR diagnostics

**DOI:** 10.1101/2021.04.01.21254755

**Authors:** Paul Naaber, Andrio Lahesaare, Laura Truu, Andres Soojärv, Ainika Adamson, Kaido Beljaev, Rainar Aamisepp, Kaspar Ratnik

## Abstract

The emerging spread of variants of concern (VOC) of SARS-CoV-2 has been noted in several countries worldwide during last months. VOCs associated with increased transmissibility and morality. Sequencing is the gold standard for investigation of variants, however it is expensive and time-consuming. S-dropout routine monitoring in combination with VOC screening by RT-PCR is a useful tool for VOC surveillance.

---

The emerging spread of variants of concern (VOC) of SARS-CoV-2 has been noted in several countries worldwide during the few last months. Such variants (e.g. B.1.1.7, B1.351, and P.1) are associated with increased transmissibility and morality, and possibly also the reduction of vaccine effectiveness [1-4]. Real-time surveillance with a prompt response is essential for the containment of their future spread.

Sequencing of positive samples is the gold standard for the typing classification of SARS-CoV-2 strains and also the identification of VOCs. However, this method is expensive and time-consuming. Although many countries have made a great effort to increase sequencing capacity it still only covers a minority of cases and the results only become available after several days or weeks. For prompt actions, large-scale surveillance with cheaper and rapid methods is required uniformly.

Recently several RT-PCR tests for the detection of SARS-CoV-2 mutations have been developed and this approach is also recommended by ECDC [5-7]. The detection of S-gene drop-out by some SARS-CoV-2 assays has also been shown as a useful tool for screening B.1.1.7.

For rapid screening of VOC, we introduced S-dropout routine monitoring in combination with VOC screening by RT-PCR in SYNLAB Estonia, Tallinn. SYNLAB is serving all of the Estonian regions and performing 79% of all Estonian SARS-CoV-2 PCR tests, so representing the whole country’s situation.

We use TaqPath COVID-19 CE-IVD RT-PCR (Thermo Fisher Scientific Inc.) for routine SARS-CoV-2 testing which is able to detect S-dropout associated with Del 69-70. S-dropouts’ counts and proportion from positive results by counties and patient groups together with trend analysis is updated daily on the COVID-19 diagnostics dashboard (Supplementary Figure).

As a pilot study, we screened 1116 positive results (a random selection from a total drawn from 1229 positive results of March 08, 2021) for Del 69-70; N501Y ja E484K by Novaplex SARS-CoV-2 Variant Assay (Seegene Inc.) to evaluate the country’s situation at the moment.

The S-dropout proportion has varied from 20 to 96% (average 78%) of all positive samples. From all of the reviewed S-dropouts, 95% were of the UK variant B.1.1.7. Thus, B.1.1.7 appeared to be the predominant genotype in Estonia, causing a total of 74% of all COVID-19 cases on this testing day. Two positive samples (0.18%) carried N501Y and E484K mutations indicating to B1.351 or P.1 variant.

For samples with N501Y and E484K mutation and randomly selected other strains amounting to 134 the sequencing was performed in the SYNLAB MVZ Humangenetik Mannheim GmbH (Germany) using commercially available Illumina COVIDSeq Test (Illumina Inc.). All UK variants (B.1.1.7) detected by RT-PCR were also confirmed by sequencing and S-dropouts without N501Y mutations belonged to lineage B.1.258. the same strain B.1.258 has been already described in the Czech Republic and Slovakia in the end of 2020 [8]. Two samples with N501Y and E484K mutations were confirmed as B.1.351 (South African variant).

## Conclusion

Continuous monitoring of S-dropouts is a useful and inexpensive tool for a follow-up of the UK variants epidemiology. However, regular cross-sectional studies for the screening of all relevant VOC should be performed since the proportion of UK strains in S-drop-outs is not stable and has significantly increased over the last 3 months in Estonia according to our data. Moreover, other important VOCs can appear and the UK strain can acquire additional mutations such as E484K.

The rapid detection of these variants means paying special attention to particular regions and patient groups as well, which should be mandatory in border SARS-CoV-2 screening, followed by prompt actions for containment of these variants.

In conclusion, RT-PCR detection of particular mutations is generally an easy and reliable way to screen thousands of samples during a short time and, together with S-dropout automated monitoring, is a useful tool for VOC containment. This approach can significantly support regular sequencing-based surveillance.

## Data Availability

Data is available by request

## References

1. Plante, J.A., Mitchell, B.M., Plante, K.S., Debbink, K., Weaver, S.C., Menachery, V.D., The Variant Gambit: COVID’s Next Move, Cell Host and Microbe (2021), doi: https://doi.org/10.1016/j.chom.2021.02.020.

2. Di Caro A, Cunha F, Petrosillo N, Beeching NJ, Ergonul O, Petersen E, Koopmans MPG. SARS-CoV-2 escape mutants and protective immunity from natural infections or immunizations. CMI 2021. https://doi.org/10.1016/j.cmi.2021.03.011

3. European Centre for Disease Prevention and Control. Risk related to spread of new SARS-CoV-2 variants of concern in the EU/EEA, first update – 21 January 2021. ECDC: Stockholm; 2021

4. Challen R, Brooks-Pollock E, Read JM, Dyson L, Tsaneva-Atanasova K, Danon L. BMJ 2021;372:579 http://dx.doi.org/10.1136/bmj.n579

5. European Centre for Disease Prevention and Control, WHO Regional Office for Europe. Methods for the detection and identification of SARS-CoV-2 variants. 3 March 2021. ECDC and WHO Regional Office for Europe: Stockholm and Copenhagen; 2021.

6. Vogels CB, Breban M, Alpert T, Petrone ME, Watkins AE, Hodcroft E, et al. PCR assay to enhance global surveillance for SARS-CoV-2 variants of concern 2021. 2021.01.28.21250486]. Available from: https://www.medrxiv.org/content/medrxiv/early/2021/02/01/2021.01.28.21250486.full.pdf.

7. Gulay Korukluoglu, Kolukirik M, Bayrakdar F, Ozgumus GG, Altas AB, Cosgun Y, et al. 40 minutes RT-qPCR Assay for Screening Spike N501Y and HV69-70del Mutations 2021. Available from: https://www.biorxiv.org/content/10.1101/2021.01.26.428302v1.full.pdf

8. Brejova, B. et al. B.1.258 Δ, a SARS-CoV-2 variant with Δ H69/ Δ V70 in the Spike protein circulating in the Czech Republic and Slovakia (2021), Bibcode: 2021arXiv210204689B.

