## Supplementary Figure for "Rapid screening for variants of concern in routine SARS-CoV-2 PCR diagnostics"

**Supplementary figures: Example figures from COVID-19 diagnostics dashboard SYNLAB Estonia**

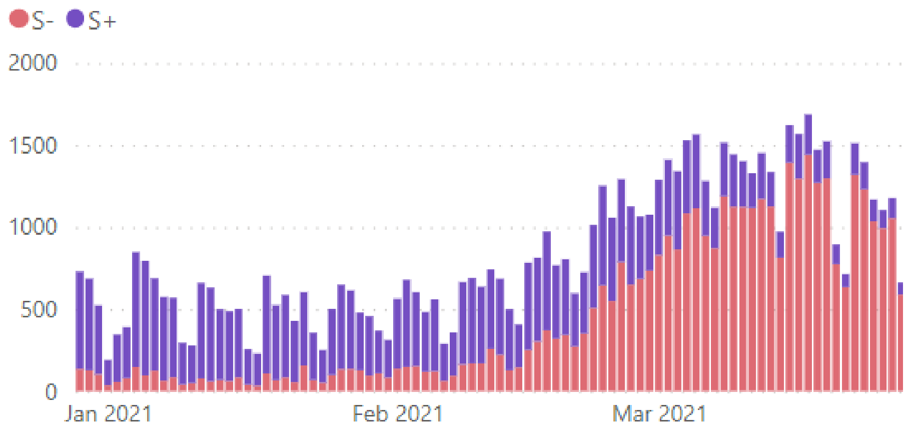


Figure 1. Positive SARS-CoV-2 PCR results per day. Red: S-dropouts; blue: non-S-dropouts

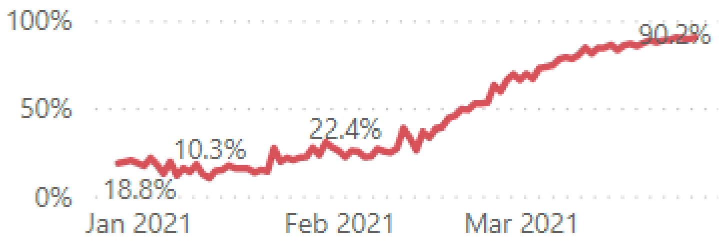


Figure 2. Changes of S-dropouts % from all positive results in time

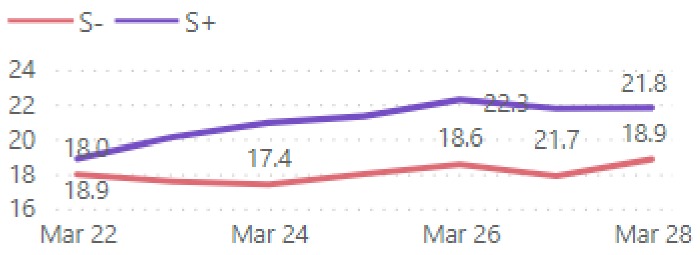


Figure 3. Differences in Ct values of N-gene. Red-S-dropouts; blue – non-S-dropouts

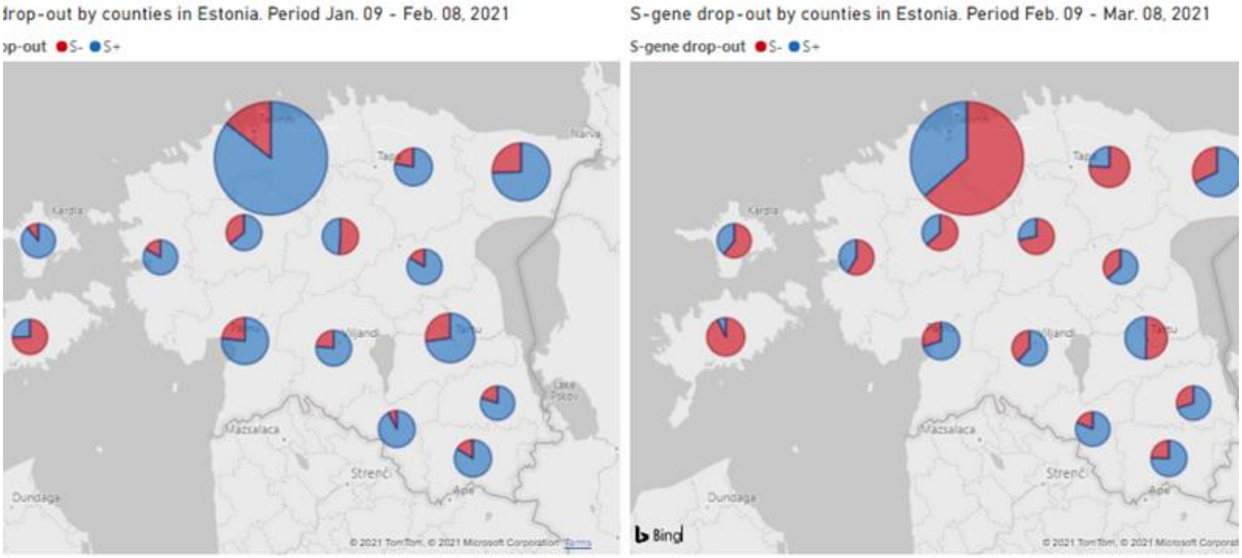


Figure 4. Number of cases (size of ring) and proportion of S-dropouts in different Estonian counties.
